# Integrating Laboratory Data and Continuous Telemonitoring to Improve Early Warning Scores

**DOI:** 10.64898/2025.12.07.25341806

**Authors:** E.E. Jerry, T.H.G.F. Bakkes, F. Schonck, R. Deneer, R.A. Bouwman, S.W. Nienhuijs

## Abstract

**Background:** Early Warning Scores (EWS) are widely implemented to identify clinical deterioration in hospitalized patients. However, their accuracy remains limited by intermittent vital sign measurements and high false-positive rates. Continuous monitoring with wearable sensors offers more timely detection, and integrating laboratory parameters may further enhance predictive precision. This study evaluated whether combining continuous vital sign monitoring with routine laboratory tests improves the prediction of postoperative complications compared with standard EWS approaches.

**Methods:** All adult patients admitted in 2023 to the surgical oncology ward of Catharina Hospital Eindhoven who underwent monitoring with a wireless accelerometer patch were included. Continuous heart and respiratory rate data were combined with laboratory parameters, including C-reactive protein (CRP) and leukocyte count. Postoperative complications (Clavien–Dindo ≥ II) were identified by chart review. Logistic regression was used to identify significant predictors and to construct the CLUE model (Combining Labdata Upgrading Early warning scores). Model performance was compared with the Continuous Remote Early Warning Score (CREWS) using the area under the receiver operating characteristic curve (AUC).

**Results:** The final dataset included 198 complete observations from 155 patients. Heart rate, CRP, and leukocyte count were independently associated with postoperative complications. The CLUE model achieved an AUC of 0.71 (0.60-0.80) outperforming the CREWS model (AUC 0.57 (0.50-0.63)).

**Conclusion:** Integrating laboratory data with continuous vital sign monitoring improved the discrimination of postoperative complications compared with vital sign based EWS alone. The CLUE model represents a clinically interpretable step toward more precise, multimodal early warning systems.

## Introduction

Early Warning Scores (EWS) are widely used tools to detect clinical deterioration. [1, 2] These systems are based on intermittent vital sign measurements, including heart rate, respiratory rate, blood pressure, and temperature, which are then scored to trigger clinical interventions. Although EWS protocols have improved the early recognition of deteriorating patients, their performance remains limited. This is due to the reliance on intermittent spot-checks, suboptimal sensitivity and specificity, and a high rate of false-positive alarms contributing to alarm fatigue. [3–5]

Continuous monitoring technologies offer an opportunity to overcome these limitations by enabling real-time detection of physiological changes. Several studies have shown that continuous tracking of heart rate and respiratory rate, the most predictive parameters of deterioration, can identify complications earlier than traditional intermittent measurements. [6–8] Wearable sensors and continuous remote EWS models, such as Continuous Remote Early Warning Score (CREWS) and Remote Early Warning Score (REWS), have demonstrated feasibility and acceptable predictive performance in surgical patients. [7, 9, 10] However, even with continuous vital sign monitoring, predictive accuracy remains to be improved.

Integrating laboratory results with vital sign monitoring may improve early detection of patient deterioration by capturing biochemical changes that precede clinical symptoms. Inflammatory markers such as C-reactive protein (CRP) and leukocyte count often rise before physiological instability becomes evident, reflecting the early stages of infection or tissue injury. [11, 12] Several studies have shown that elevated postoperative CRP or leukocyte levels are associated with an increased risk of complications and prolonged recovery after major surgery. [13, 14] Combining these laboratory trends with continuous vital sign data may therefore provide a more comprehensive and timely assessment of patient condition than vital signs alone.

The present study aimed to evaluate whether combining continuous vital sign monitoring with routinely collected laboratory parameters could improve the prediction of postoperative complications compared with standard vital sign based EWS models. This approach may support earlier identification of high-risk patients while reducing false-positive alerts and alarm burden in surgical wards. Ultimately supporting more efficient workflows in acute care settings.

## Methods

### Ethics statement

Ethical approval for this study was obtained from the Medical Research Ethics Committee (MEC-U) and hospital’s local ethics committee with the reference number NCT05956210.

### Study design and setting

This single-center retrospective cohort study was conducted at Catharina Hospital Eindhoven, a large teaching hospital in the Netherlands. The ward primarily admits patients undergoing major abdominal oncological procedures such as colorectal, esophageal, gastric, bladder, and gynecological surgery.

### Participants and data collection

The study population comprised all adult patients admitted in 2023 to the surgical oncology ward who were monitored using a wireless accelerometer patch for telemonitoring (Healthdot®, Philips, Eindhoven, the Netherlands). Patients were eligible if Healthdot data were available during their admission. From this population, patients with available laboratory data were included. Complications were classified and graded according to the Clavien–Dindo classification system. [15] For the analyses, complications of grade II or higher were considered clinically relevant adverse outcomes. All routinely collected laboratory parameters were extracted from the electronic medical record, including hematology, chemistry, coagulation, and point-of-care (POC) glucose measurements. These comprised markers such as CRP, leukocyte count, hemoglobin, hematocrit, electrolytes, renal and liver function tests, coagulation parameters, and urinary screening results. All available laboratory values were temporally aligned with continuous vital sign data from the wearable device.

### Wearable device and data processing

The Healthdot is a small wireless accelerometer-based sensor applied on the left thoracic wall. It continuously records heart rate and respiratory rate. Every 5 minutes, averages of the collected data over the past 5 minutes are automatically transmitted via a low-power LoRa (Long Range) network to a secure hospital database. The device has been validated for accuracy in surgical patients. [16] Because wearable and laboratory data differ substantially in sampling frequency, a structured temporal alignment process was applied.

### Data Engineering and model development

Routine laboratory tests were generally performed between 07:00 and 10:00, with occasional additional measurements throughout the day. Each postoperative complication or discharge event was treated as a distinct observation in the dataset. For every observation, the most recent laboratory measurement obtained within a 48-to 6-hour window prior to the event was selected and linked to that observation. The 6-hour buffer period ensured that only data preceding the event were used for analysis, preventing label leakage.

Laboratory measurements were used as anchor points for temporal alignment of wearable data. Based on the timestamp of each selected laboratory result, continuous data from the Healthdot wearable (heart rate and respiratory rate) were aggregated over the preceding four-hour period. The median values of these parameters were calculated and assigned to the corresponding laboratory observation. This four-hour aggregation window corresponded to the temporal resolution applied in the CREWS, allowing a direct methodological comparison between the two approaches. To ensure completeness and consistency, all observations with missing values for any of the included variables were excluded from the analysis. In Table 1, the Continuous Remote Early Warning Score (CREWS) is calculated based on the work by Van Der Stam et al. [10] which utilizes heart rate and respiration rate to assign scores based on specific thresholds.

**Table 1.** CREWS.

| Vital Parameter | Score |  |  |
| --- | --- | --- | --- |
|  | 0 | 1 | 2 |
| Heart Rate (bpm) | 40 - 110 |  | <40 or > 110 |
| Respiration Rate (bpm) | 8 - 20 | <8 or > 20 |  |

A univariate Mann-Whitney U-test was used to assess the association between physiological and laboratory parameters and postoperative complications (Clavien–Dindo grade ≥ II). All variables were subsequently entered into a logistic regression model using a forward selection procedure. A model was initiated with only the heart rate as parameter. Subsequently, additional predictors were added iteratively based on their ability to reduce the mean squared error and the process was continued until the model comprised three parameters. This stepwise procedure was used to force the model to start with heart rate as the initial parameter and to limit the number of parameters in the model to three. Thereby, keeping the model simple and easier to evaluate with a limited number of additional lab tests. Heart rate was chosen as the starting parameter as it had the highest predictive value of the two continuous parameters available from the Healthdot.

To facilitate clinical interpretation, the regression coefficients were converted into a quantized scoring system. Two fitted-value thresholds, corresponding to model performance at approximately 0.5 specificity and 0.1 sensitivity, were used to define clinically meaningful transitions in predicted risk. Each variable was divided into three levels (0, 1, and 2), consistent with the design of established early warning scores such as NEWS, MEWS, and CREWS. The variable-specific cutoff values were determined by calculating the deviation from the mean required to achieve the predefined change in model output, adjusted for the relative contribution of each coefficient. The resulting quantized model was referred to as the CLUE score.

### Statistical analysis

Statistical analyses were performed using Python (version 3.12; Python Software Foundation) and standard scientific libraries (pandas, NumPy, and scikit-learn). Continuous variables were reported as medians with interquartile ranges (IQRs) and compared using Mann-Whitney U-tests. The process of constructing the constructing multivariate logistic regression model and the CLUE score derived from it was repeated 10.000 times using bootstrapping. The discriminative performance of the CLUE score and the CREWS was evaluated using the out of bag samples of each iteration. The Area under the receiver operating characteristics curve (AUC), confusion matrices, and calibration plots were generated to assess classification accuracy, sensitivity, and specificity.

## Results

The data collection process yielded a total of 338 observations from 250 unique patients. After excluding observations with missing values to ensure data completeness, the final dataset consisted of 198 observations from 155 patients.

To assess the individual predictive value of each parameter, a univariate analysis was performed using the Mann-Whitney U-test. The resulting p-values are presented in Table 2, with statistically significant parameters highlighted in bold.

**Table 2:** P-value resulting from the univariate Mann-Whitney U-test.

| Parameter | p-value | Parameter | p-value |
| --- | --- | --- | --- |
| RF | **0.004** | MCV | 0.250 |
| HR | **<0.001** | MCHC | 0.068 |
| **Hemoglobin** | **<0.001** | RDW.SD | 0.056 |
| **Hematocrit** | **<0.001** | **Erythroblasts** | **<0.001** |
| Thrombocytes | 0.130 | **Leukocytes** | **<0.001** |
| **Erythrocytes** | **<0.001** | **CRP** | **<0.001** |
| MCH | 0.110 |  |  |

Significant parameters ( $p < 0.05$ ) have been labelled bold. RF = respiration frequency, HR = heart rate, CRP = C-reactive protein, MCV = mean corpuscular volume, MCH = mean corpuscular hemoglobin, MCHC = mean corpuscular hemoglobin concentration, RDW.SD = Red Cell Distribution Width. Standard Deviation

Subsequently, a multivariate logistic regression model was fitted over 10000 iterations using bootstrapping. The distributions of the coefficients of these models over the iterations are shown in Fig. 1. The calibration plot of the model can be found in Fig. 2.

**Figure 1:**
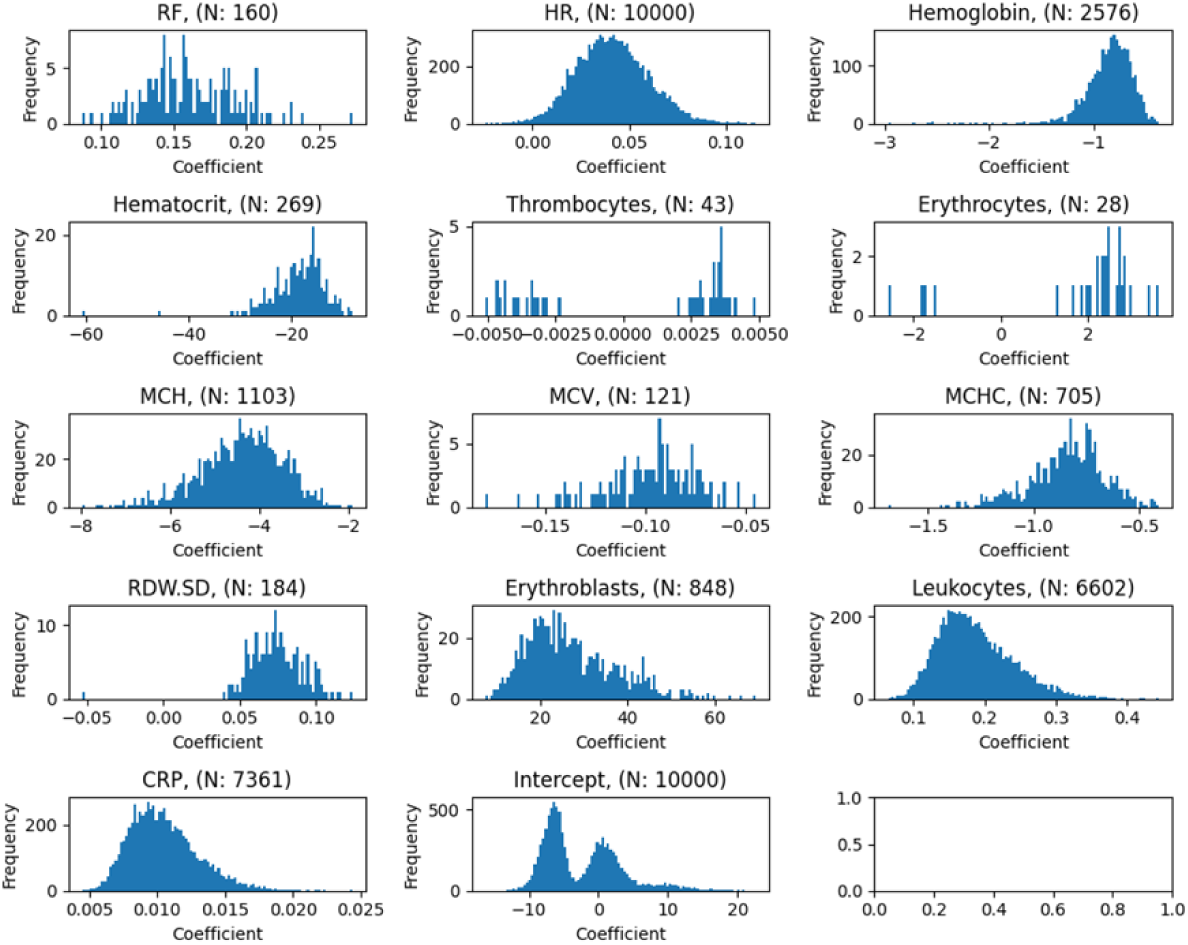
Parameter coefficients of multivariate model The coefficient distribution over the different bootstrap iterations are show. The N number indicates in how many iteration the variable was selected.

**Figure 2:**
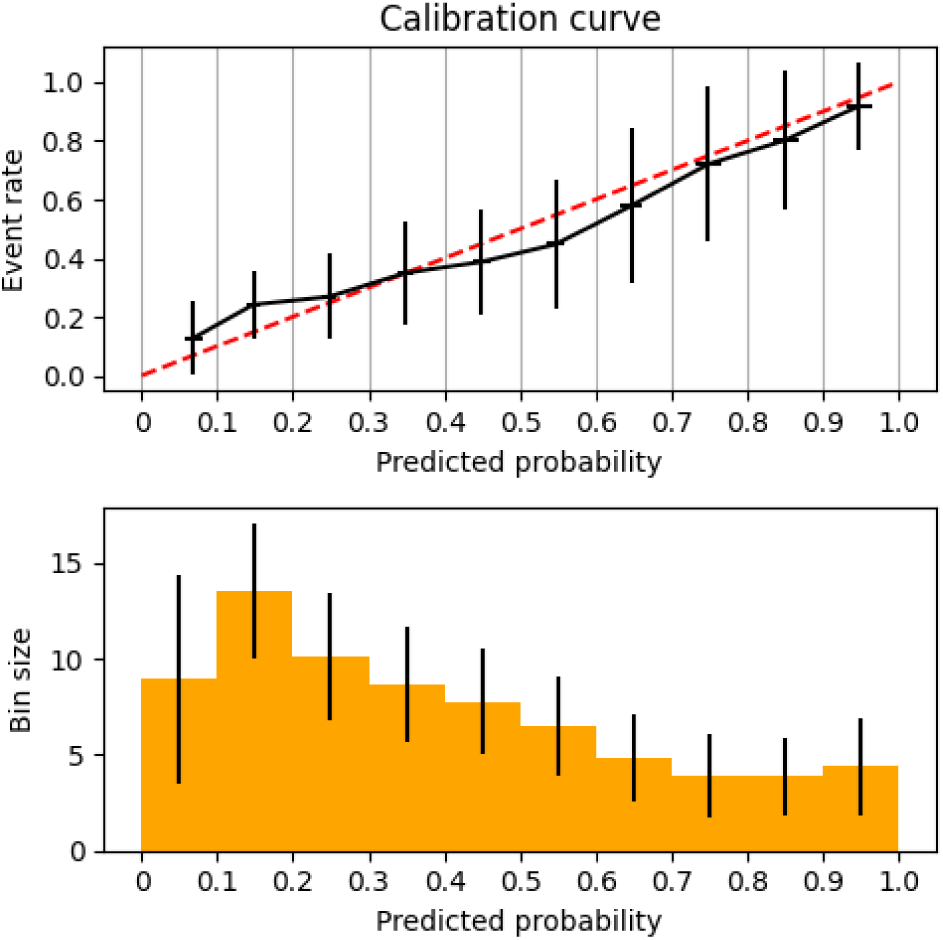
Calibration plot The top panel shows the average calibration curve across 10 bins, with black lines indicating the standard deviation. The bottom panel presents a bar chart of the average bin sizes along with their corresponding standard deviations.

The multivariate model was transformed into a quantized scoring system. This system discretizes each parameter into three levels, facilitating clinical interpretation and comparison with established scores.

During the bootstrapping the multivariate model is refitted during each iteration. Thus, resulting in a different combination of parameters each time. To visualize the construction of the CLUE score the 3 most common occurring combinations of parameters are visualized in Figure 3. The coefficients used to construct this plot are based on the median coefficient value of this combination of parameters. The three resulting CLUE scores can be found in table 4.

**Figure 3:**
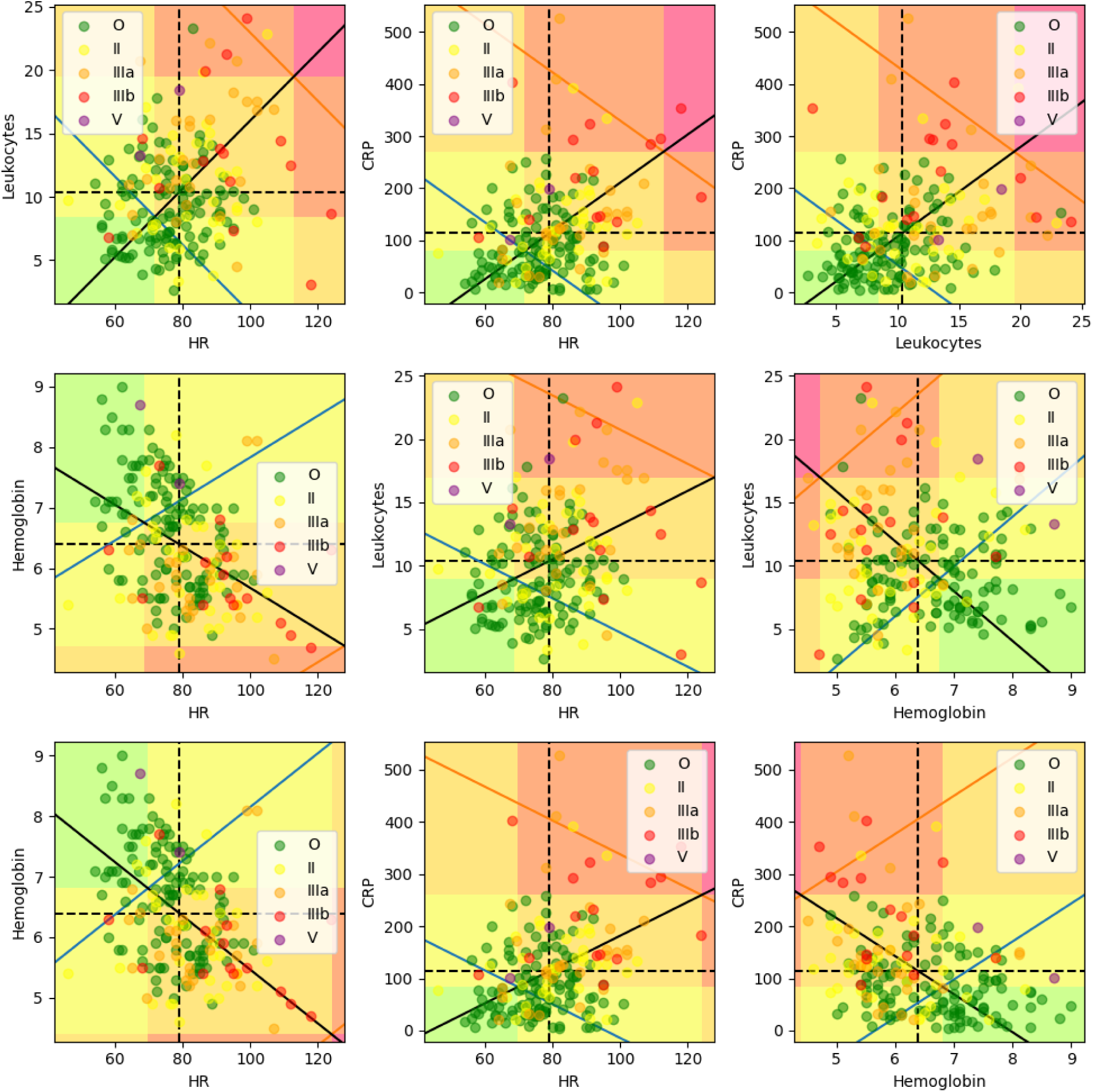
Visualization of all pairwise combinations All pairwise combinations of the three model parameters of the three most common combinations of parameters. Each dot represents a patient observation, color-coded by Clavien-Dindo classification. The dotted lines indicate the mean values of each parameter. The solid black line represents the direction in parameter space along which all three variables contribute equally to the model’s predicted outcome, based on the multivariate coefficients. The blue and orange lines mark the fitted value thresholds corresponding to the transitions from score level 0 to 1 and from 1 to 2, respectively. The intersection points between these thresholds and the equal-contribution line determine the parameter-specific cut-offs used in the scoring system.

**Table 4:** The transition values between the different levels for the CLUE score.

| Model 1 | 0 | 1 | 2 |
| --- | --- | --- | --- |
| HR | > 71.8 | $\leq 71.8$ and > 113 | $\leq 113$ |
| Leukocytes | < 8.43 | $\geq 8.43$ and < 19.5 | $\geq 19.5$ |
| CRP | < 80.3 | $\geq 80.3$ and < 270 | $\geq 270$ |
| Model 2 | 0 | 1 | 2 |
| HR | < 68.6 | $\geq 68.6$ and < 128 | $\geq 128$ |
| Hemoglobin | > 6.75 | $\leq 6.75$ and > 4.72 | $\leq 4.72$ |
| Leukocytes | < 8.97 | $\geq 8.97$ and < 17.0 | $\geq 17.0$ |
| Model 2 | 0 | 1 | 2 |
| HR | < 69.8 | $\geq 69.8$ and < 124 | $\geq 124$ |
| Hemoglobin | > 6.81 | $\leq 6.81$ and > 4.39 | $\leq 4.39$ |
| CRP | < 83.8 | $\geq 83.8$ and < 260 | $\geq 260$ |

Model performance was evaluated using the Area Under the Receiver Operating Characteristic curve (AUROC). In addition, the newly developed score was compared to existing scoring systems, specifically the CREWS and CLUE scores, using confusion matrices. The AUROC curves are presented in Figure 4, while the average confusion matrices for each score are shown in Tables 5 and 6, respectively.

**Fig. 4:**
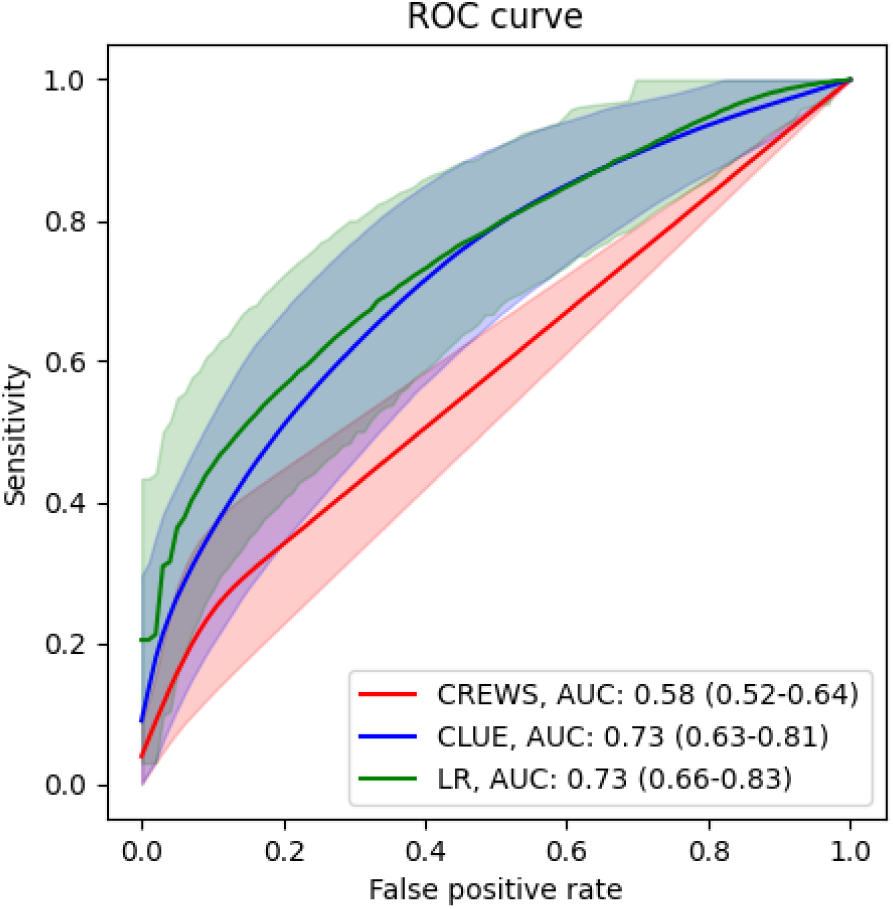
Receiver operating characteristics curves comparing model performance Shaded area indicates the 5% to 95% interval over all the iterations of the bootstrapping.

**Table 5:** Average confusion matrix for CREWS score.

| CREWS<br>Clavien<br>Dindo Score | 0 | 1 | 2 | 3 |
| --- | --- | --- | --- | --- |
| O | 38.9 | 4.76 | 0.00 | 0.00 |
| II | 11.7 | 1.09 | 0.00 | 0.00 |
| IIIa | 6.25 | 3.67 | 0.00 | 0.00 |
| IIIb | 2.56 | 1.84 | 0.37 | 0.74 |
| V | 0.73 | 0.00 | 0.00 | 0.00 |

**Table 6:** Average confusion matrix for CLUE score.

| CLUE<br>Clavien<br>Dindo Score | 0 | 1 | 2 | 3 | 4 | 5 | 6 |
| --- | --- | --- | --- | --- | --- | --- | --- |
| O | 6.50 | 14.4 | 13.1 | 8.15 | 1.34 | 0.13 | 0.00 |
| II | 0.72 | 2.33 | 4.81 | 3.55 | 1.06 | 0.36 | 0.00 |
| IIIa | 0.52 | 0.89 | 2.38 | 3.31 | 2.38 | 0.40 | 0.04 |
| IIIb | 0.03 | 0.49 | 0.81 | 1.59 | 1.92 | 0.60 | 0.07 |
| V | 0.01 | 0.16 | 0.29 | 0.16 | 0.10 | 0.00 | 0.00 |

## Discussion

This study demonstrates that integration of continuous vital sign monitoring with routinely collected laboratory parameters enhances the prediction of postoperative complications. The model, combining heart rate, leukocyte count, and CRP, outperformed the Continuous Remote Early Warning Score in discriminative ability, indicating that biochemical and physiological data provide complementary insights into patient status. Traditional EWS rely exclusively on intermittently measured vital signs, which limits temporal resolution and sensitivity to early physiological changes. [1, 2, 4, 5] Although continuous monitoring systems using wearable sensors have shown promising results in improving the timeliness of deterioration detection [6–8, 10], their predictive accuracy remains modest, with reported AUC values typically between 0.65 and 0.75. [4, 7] This study adds to a growing body of evidence that multimodal data integration, specifically combining laboratory and continuous physiological data, can substantially improve predictive performance.

While previous research has demonstrated the utility of combining laboratory parameters with EWS, the novelty of this study lies in the integration of high-frequency continuous monitoring with routine laboratory tests. Similar approaches, such as combining biomarkers with the MEWS, monocyte distribution width, and neutrophil-to-lymphocyte ratio [17], as well as integrating EWS with scores like the SOFA score (which includes laboratory data), have been explored in the literature. For example, Chen et al. (2025) used machine learning methods with routine laboratory parameters to predict bloodstream infections in ICU patients. [18] These studies highlight the increasing interest in incorporating laboratory data into EWS frameworks, reinforcing the importance of multimodal data in early detection systems.

Moreover, the use of a limited number of robust parameters (heart rate, CRP, and leukocytes) demonstrates that even simple multimodal models can outperform purely vital sign–based systems. This parsimonious design enhances interpretability and implementation feasibility, which are critical for clinical adoption. While many machine learning–based EWS models report higher predictive accuracy, their “black box” nature and dependence on large-scale data integration often limit transparency and clinical trust. [19, 20] The CLUE model, by contrast, balances predictive performance with interpretability, making it suitable for integration in clinical workflows or bedside dashboards.

From a clinical perspective, the integration of laboratory and wearable data could support more dynamic and individualized monitoring strategies. Inflammatory markers and early metabolic changes may manifest before visible physiological deterioration. This could enable earlier intervention and more accurate diagnostic reassessment. Additionally, a multimodal approach could help reduce false alarms [2, 3], which are a significant contributor to alarm fatigue in remote monitoring systems. Although future research should investigate the real-time integration of multimodal monitoring into EHR systems, immediate actions are already feasible.

Several limitations should be acknowledged. The study employed a single-center retrospective design with a moderate sample size, which may limit the generalizability of the findings. A potential source of bias arises from the exclusion of observations with incomplete data, which could affect the robustness of the model. Additionally, laboratory tests were performed intermittently, typically once daily, which may limit the temporal granularity compared to the continuous data stream provided by the wearable sensors. The current clinical practice already heavily relies on laboratory results for clinical decision-making. For example, increased CRP levels frequently lead to interventions such as CT imaging and surgical procedures, both of which are accounted for in the Clavien-Dindo classification applied in this study. This clinical practice bias could influence the observed outcomes, as laboratory data may already be shaping clinical decisions before the wearable data is even analyzed. Although external validation is indispensable for large-scale deployment, internal validation within the study cohort is equally critical. Such validation would offer a rigorous assessment of the CLUE model’s predictive performance in real-world clinical practice, thereby enabling refinements that enhance both accuracy and generalizability before broader implementation.

Despite these limitations, the study is strengthened by its integration of high-frequency wearable data with routinely collected laboratory results in a real-world context, supported by a transparent data engineering framework that maintains temporal integrity. The structured temporal alignment of wearable and laboratory data represents an innovative step toward multimodal predictive models. The quantized, interpretable format of the CLUE model facilitates clinical adoption by translating complex statistical outputs into actionable insights. A central methodological strength of this study is the data-engineering framework, which aligns laboratory timestamps and retrospectively aggregates wearable data within physiologically meaningful windows. This mitigates the risk of label leakage and allows clinically meaningful synchronization between continuous and episodic data sources. Similar strategies are increasingly employed in multimodal clinical prediction research [21–23], yet few have been implemented or validated in real-world postoperative monitoring contexts. By discretizing model coefficients into a quantized, interpretable score, CLUE translates statistical complexity into a clinically usable format, aligning with calls for transparent and explainable predictive systems in healthcare. [24]

In summary, the CLUE model offers a transparent methodological framework with clear clinical interpretability, integrating continuous physiological monitoring and biochemical markers to improve early detection of postoperative complications. By linking laboratory and wearable data, it outlines a viable route toward multimodal early warning systems that balance interpretability, predictive performance, and clinical applicability.

## Data Availability

The data underlying this study will be made fully available upon request. Please contact the corresponding author for access.

## References

1. Smith, G.B., et al., The National Early Warning Score 2 (NEWS2). (1473-4893 (Electronic)).

2. Gerry, S., et al., Early warning scores for detecting deterioration in adult hospital patients: a systematic review protocol. (2044-6055 (Electronic)).

3. Smith, G.B., et al., The ability of the National Early Warning Score (NEWS) to discriminate patients at risk of early cardiac arrest, unanticipated intensive care unit admission, and death. (1873-1570 (Electronic)).

4. Cvach, M., Monitor alarm fatigue: an integrative review. (0899-8205 (Print)).

5. Sendelbach, S. and M. Funk, Alarm fatigue: a patient safety concern. (1559-7776 (Electronic)).

6. Taenzer, A.H., S.P. Pyke Jb Fau - McGrath, and S.P. McGrath, A review of current and emerging approaches to address failure-to-rescue. (1528-1175 (Electronic)).

7. van Ede, E.S., et al., Continuous remote monitoring in post-bariatric surgery patients: development of an early warning protocol. (1878-7533 (Electronic)).

8. Weenk, M.A.-O., et al., Continuous Monitoring of Vital Signs Using Wearable Devices on the General Ward: Pilot Study. (2291-5222 (Print)).

9. Pimentel, M.A.F., et al., A comparison of the ability of the National Early Warning Score and the National Early Warning Score 2 to identify patients at risk of in-hospital mortality: A multi-centre database study. (1873-1570 (Electronic)).

10. van der Stam, J.A., et al., A wearable patch based remote early warning score (REWS) in major abdominal cancer surgery patients. (1532-2157 (Electronic)).

11. Neumaier, M. and M.A. Scherer, C-reactive protein levels for early detection of postoperative infection after fracture surgery in 787 patients. (1745-3682 (Electronic)).

12. Warschkow R Fau - Steffen, T., et al., Diagnostic accuracy of C-reactive protein and white blood cell counts in the early detection of inflammatory complications after open resection of colorectal cancer: a retrospective study of 1,187 patients. (1432-1262 (Electronic)).

13. Welsch, T., et al., C-reactive protein as early predictor for infectious postoperative complications in rectal surgery. (0179-1958 (Print)).

14. Ortega-Deballon, P., et al., C-reactive protein is an early predictor of septic complications after elective colorectal surgery. (1432-2323 (Electronic)).

15. Bolliger, M., et al., Experiences with the standardized classification of surgical complications (Clavien-Dindo) in general surgery patients. (1682-8631 (Print)).

16. van der Stam, J.A., et al., Accuracy of vital parameters measured by a wearable patch following major abdominal cancer surgery. (1532-2157 (Electronic)).

17. Lin, S.A.-O., et al., A novel scoring system combining Modified Early Warning Score with biomarkers of monocyte distribution width, white blood cell counts, and neutrophil-to-lymphocyte ratio to improve early sepsis prediction in older adults. (1437-4331 (Electronic)).

18. Chen, Y., et al., Early prediction of bloodstream infections in ICU patients using machine learning methods based on routine laboratory parameters. (1471-2334 (Electronic)).

19. Ghassemi, M., et al., A Review of Challenges and Opportunities in Machine Learning for Health. (2153-4063 (Print)).

20. Rudin, C., Stop Explaining Black Box Machine Learning Models for High Stakes Decisions and Use Interpretable Models Instead. (2522-5839 (Electronic)).

21. Escobar, G.J., et al., Risk-adjusting hospital mortality using a comprehensive electronic record in an integrated health care delivery system. (1537-1948 (Electronic)).

22. Churpek, M.M., et al., Multicenter development and validation of a risk stratification tool for ward patients. (1535-4970 (Electronic)).

23. Thorsen-Meyer, H.C., et al., Dynamic and explainable machine learning prediction of mortality in patients in the intensive care unit: a retrospective study of high-frequency data in electronic patient records. (2589-7500 (Electronic)).

24. Johnson, A.E.W., et al., MIMIC-IV, a freely accessible electronic health record dataset. (2052-4463 (Electronic)).

